# Broad phenotypic alterations and potential dysfunctions of lymphocytes in COVID-19 recovered individuals

**DOI:** 10.1101/2020.07.01.20144030

**Authors:** Jingyi Yang, Maohua Zhong, Ejuan Zhang, Ke Hong, Qingyu Yang, Dihan Zhou, Jianbo Xia, Yao-Qing Chen, Mingbo Sun, Bali Zhao, Jie Xiang, Ying Liu, Yang Han, Xi Zhou, Chaolin Huang, You Shang, Huimin Yan

**Author notes:** Contributed equally (Jingyi Yang, Maohua Zhong, Ejuan Zhang, Ke Hong). Correspondence (H.Y.), (Y.S.), (C.H.), (X.Z.).

## Abstract

**Background:** Lymphopenia is a typical symptom in the COVID-19 patients. While millions of patients are clinical recovered, little is known about the immune status of lymphocytes in these individuals.

**Methods:** A clinical recovered cohort (CR) of 55 COVID-19 individuals (discharged from hospital 4 to 11 weeks), and 55 age and sex matched healthy donors cohort (HD) were recruited. Detailed analysis on phenotype of the lymphocytes in peripheral blood mononuclear cells (PBMCs) was performed by flow cytometry.

**Findings:** Compared with cohort HD, the CD8^+^ T cells in cohort CR had higher Teff and Tem, but lower Tc1 (IFN-γ^+^), Tc2 (IL-4^+^) and Tc17 (IL-17A^+^) frequencies. The CD4^+^ T cells of CR had decreased frequency, especially on the Tcm subset. Moreover, CD4^+^ T cells of CR expressed lower PD-1 and had lower frequencies of Th1 (IFN-γ^+^), Th2 (IL-4^+^), Th17 (IL-17A^+^) as well as circulating Tfh (CXCR5^+^PD-1^+^). Accordingly, isotype-switched memory B cell (IgM^-^CD20^hi^) in CR had significantly lower proportion in B cells, though level of activation marker CD71 elevated. For CD3^-^HLA-DR^lo^ lymphocytes of CR, besides levels of IFN-γ, Granzyme B and T-bet were lower, the correlation between T-bet and IFN-γ became irrelevant. In addition, taken into account of discharged days, all the lowered function associated phenotypes showed no recovery tendency within whole observation period.

**Interpretation:** The CR COVID-19 individuals still showed remarkable phenotypic alterations in lymphocytes after clinical recovery 4 to 11 weeks. This suggests SARS-CoV-2 infection imprints profoundly on lymphocytes and results in long-lasting potential dysfunctions.

**Funding:** Kunming Science and Technology Department (2020-1-N-037)

## Introduction

The worldwide pandemic of COVID-19 caused by SARS-CoV-2 has become global threat to human and society though the epidemic subsided after control measures were taken in China. As of Jun 30, 2020, there were more than 10 million confirmed infections globally, in more than 180 countries, with more than 500,000 deaths, and about 5.6 million clinical recovered. Lymphocytes are critical for eliminating infection and for establishment of long-term immunity.^1-3^ However, lymphopenia was reported as a typical clinical symptom in COVID-19 patients besides the specific peripheral ground-glass opacities lung consolidation.^4,5^ A growing of studies have reported decrease number and impaired function of CD4^+^ T, CD8^+^ T and NK cells in COVID-19 patients, especially in severe COVID-19 cases.^6-11^ Although lymphocyte count could gradually elevate to normal range in some patients in blood after viral clearance and clinical recovery,^7,12^ the activation, differentiation and function of lymphocytes in the clinical recovered COVID-19 subjects remain poorly understood. In a follow-up study of an asymptomatic infected case without lymphopenia, we found IFN-γ^+^CD8^+^ T cells and IL-17A^+^CD4^+^ T cells were still detained at a remarkable lower percentages, even when SARS-CoV-2 virus had become undetectable for three weeks.^13^ These results whistle that SARS-CoV-2 may persistently affect the immune system. Despite the large numbers of recovered populations, there is a scarcity of information on the composition, phenotype and function potential of the lymphocytes. In this study, we comprehensively investigated the lymphocyte phenotype and potential function of a clinical recovered COVID-19 cohort (CR) recruited in Wuhan by comparative analyzing the peripheral blood nonnuclear cells (PBMCs) with age & sex matched uninfected healthy donors (HD).

## Methods

### Study design and participants

We did a cohort study on individuals aged 25-70 years in Wuhan Jinyintan Hospital in April 2020. We collected peripheral blood nonnuclear cells (PBMCs) from 72 COVID-19 recovered individuals who were recruited for physical reexamination after discharge at least 4 weeks, and from 57 healthy donors control who were recruited for regular physical examination.

Clinical recovered COVID-19 individuals with virus re-detected positive (3 persons), and individuals with both SARS-CoV-2 RBD specific IgM and IgG negative in plasma (13 persons), were excluded. Healthy donors were enrolled and confirmed without on-going or past SARS-CoV-2 infection, based on nucleotide acid assay of nasopharyngeal swab samples and SARS-CoV-2 RBD specific IgM and IgG assays of plasma (2 persons excluded). The individuals who had series chronic problem such as HIV, HCV infection, cerebrovascular disease, kidney disease were also excluded (1 Clinical recovered COVID-19 individual).

After exclusion, a cohort of fifty-five healthy donors (HD, 23 male and 32 female, mean age 49·1, median age 51) and a fifty-five clinical recovered COVID-19 cohort (CR, 21 male and 34 female, mean age 48·8, median age 51) were involved in the study (supplement table 1), except the cases for assay on Tfh, B cells and cytokines IL-2, in which a sub-cohort of only 36 clinical recovered individuals (14 male and 22 female, mean age 50·6, median age 52) was involved in due to lack of antibody reagents.

This study was reviewed and approved by the Medical Ethical Committee of Wuhan Jinyintan hospital (approval number KY-2020-47·01). Written informed consent was obtained from the COVID-19 recovered individuals and the healthy donors.

### Clinical laboratory measurements

Clinical laboratory measurements, antibody detection were performed in Wuhan Jinyintan Hospital in April, 2020. Nasopharyngeal swab samples were collected on the day of peripheral blood collection and were tested by qRT-PCR for amplification of E gene, RdRp gene, and N gene of SARS-CoV-2 as described by our previous study. ^14^

Clinical laboratory investigation was performed, including series of complete blood count, physical examination and etc (supplementary table 1).

### Lymphocyte responses evaluation by flow cytometry

Plasma and cell pellets were separated from fresh peripheral blood from the clinical recovered COVID-19 individuals and healthy donors. Plasma was used for detection of SARS-CoV-2 binding IgM or IgG. PBMCs were separated from the cell pellets after resuspension with PBS by density gradient centrifugation and resuspended in complete RPMI1640 medium containing 10% FBS (Gibco), 1% penicillin and 1% streptomycin.

For phenotypical analysis of lymphocytes, PBMCs were staining directly. For functional assay of lymphocytes, PBMCs were stimulated with 200 ng/ml PMA (Beyotime, China), 2·5 μM ionomycin (Beyotime, China) in the presence of 1 μM monensin (BioLegend, USA) and 2·5 μg/ml Brefeldin A at 37°C, 5% CO_2_ for 4·5 h before staining. PBMCs were stained with dead cell discrimination marker (eBioscience™ Fixable Viability Dye eFluor™ 506) and a panel of surface mAbs in PBS at 4°C for 30 min. After washed by PBS, cells were fixed with fixation/permeabilization buffer (eBioscience) at 4°C overnight, and then stained with the respective panel of intracellular markers in a permeabilization buffer at 4°C for 30 min. Antibodies used in this study were all from BioLegend (USA) and listed in supplementary table S2. A BD LSR Fortessa flow cytometer (Becton Dickinson) was used to assess the stained cells. Data was analyzed using FlowJo V7.0.

### Statistical analysis

Data are presented as means±SD. All data analyses were performed with unpaired 2 -tailed Student’s t-test or Liner Regression. Statistical analysis was carried out with InStat, version 8·0 (GraphPad Software, La Jolla, CA, USA). P < 0·05 was considered significant.

### Role of the funding source

The funding agencies did not participate in study design, data collection, data analysis, or writing of the report. The corresponding authors were responsible for all aspects of the study to ensure that issues related to the accuracy or integrity of any part of the work were properly investigated and resolved. The final version was approved by all authors.

## Results

On the day of sample collection and assay, all subjects of CR were in normal oxygen saturation over 95% by clinical examination (Supplementary Table S1). The majority of CR individuals presented no symptoms in the past two weeks, except four experienced shod of breath, six experienced cough, three experienced expectoration, and one experienced oxygen uptake.

All the 55 CR individuals had normal counts of total lymphocytes, except one showed remarkable lymphopenia (0·64*10^7^/ml) (supplementary table S1). Lymphocyte populations, including CD8^+^ T cells, CD4^+^ T cells, CD3^-^CD19^+^ B cells, and non-T non-B CD3^-^HLA-DR^lo^ subset were analyzed (figure 1A). To our surprise, percentage of CD4^+^ T cells in the lymphocytes of CR cohort was statistically lower than that in the healthy donors (HD), though the percentage of CD8^+^ T cells, B cells or CD3^-^HLA-DR^lo^ lymphocytes in the lymphocytes of CR cohort was already comparable to that of HD (figure 1B). We then assayed and analyzed the differentiation, activation, proliferation and potential function of the four lymphocyte subsets one by one.

**Figure 1.**
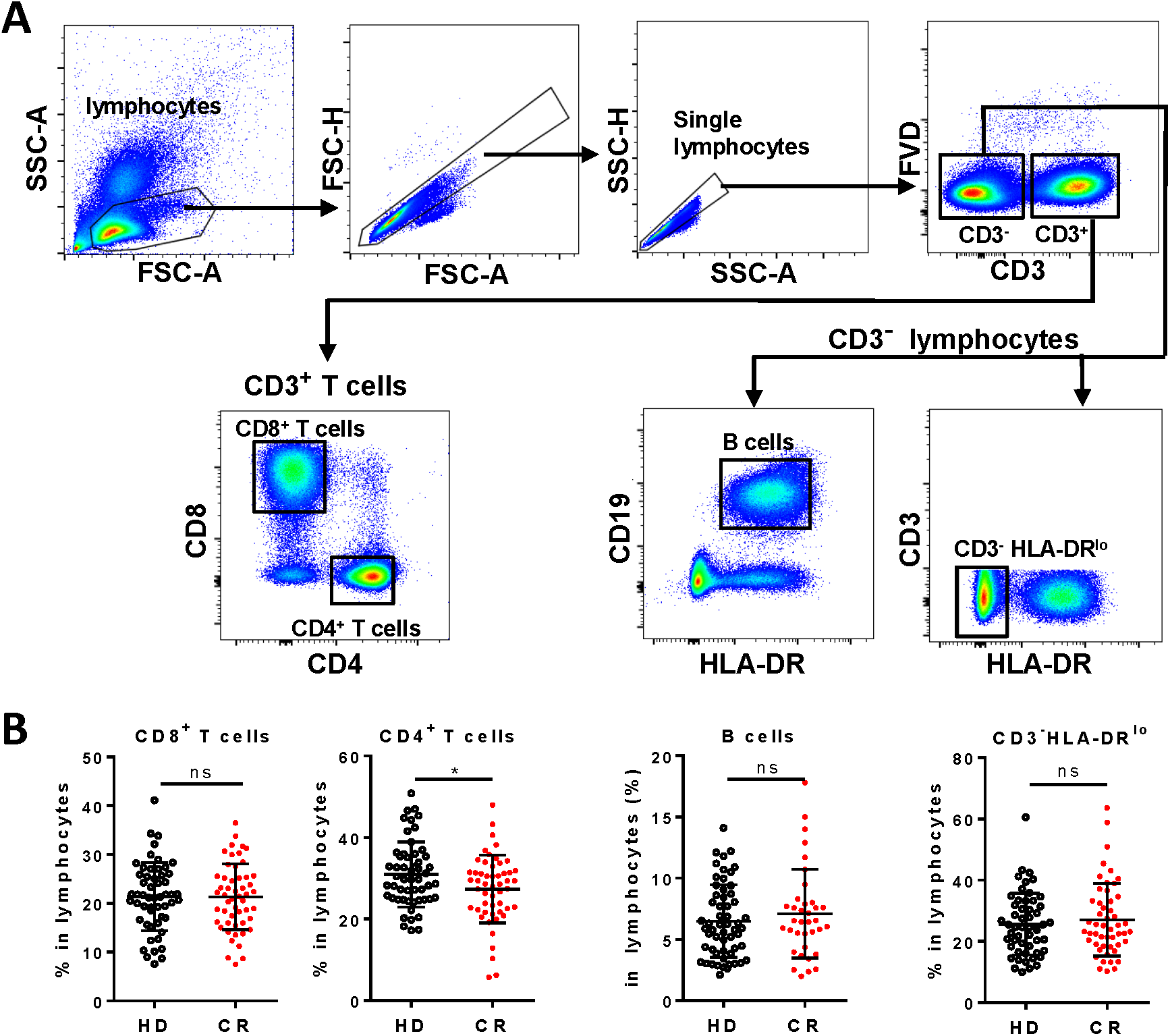
CD8^+^ T cells, CD4^+^ T cells, B cells and CD3^-^HLA-DR^lo^ lymphocytes in peripheral blood of healthy donor cohort (HD) and clinical recovered cohort (CR). A, Gating strategies of CD8^+^ T cells (CD3^+^CD4^-^CD8^+^), CD4^+^ T cells (CD3^+^CD8^-^CD4^+^), B cells (CD3^-^CD19^+^ HLA-DR^+^) and CD3-HLA-DR^lo^ lymphocytes (CD3^-^HLA-DR^lo^) in PBMCs. B, Frequencies of CD8^+^ T cells, CD4^+^ T cells, B cells and CD3-HLA-DR^lo^ lymphocytes in lymphocytes of HD and CR. ns, non-significant; *, P<0.05.

In the context of CD8^+^ T cells, the frequencies of both effector T cells (CD45RO^-^CD27^-^, Teff) and effector memory T cells (CD45RO^+^CD27^-^, Tem) of CR were significantly higher than that of HD, while the frequency of CR naïve T cells (CD45RO ^-^CD27^+^, Tna) and central memory T cells (CD45RO^+^CD27^+^, Tcm) were lower than that of HD (figure 2A). This indicated significant differentiation occurred in CD8^+^ T cells post SARS-CoV-2 infection. In terms of activation, HLA-DR and PD-1 of CR cohort were similar as those of HD (figure 2B and S1A, S1B), indicating CD8^+^ T cells in CR nearly recovered into normal level, although with a slightly higher proliferation activity indicated by Ki-67^+^ in Tcm subsets (figure 2C and S1C). Then, functions of CD8^+^ T cells were analyzed under polyclonal stimulation. Intriguingly, although frequencies of IL-2^+^ and Granzyme B (GZMB)^+^ CD8^+^T cells showed no difference between CR and HD (figure 2D), the frequency of IFN-γ^+^ in CD8^+^ T cells (Tc1) of CR was notably reduced compared with that of HD (figure 2E). However, T-bet expression on CD8^+^ T cells of CR was not reduced and the correlation between T-bet^+^ and IFN-γ^+^ remained (figure 2F). Further analysis showed that proportions of IFN-γ^+^IL-2^-^ and IFN-γ^+^GZMB^-^ in CR cohort were significantly reduced, while IFN-γ^-^GZMB^+^ remarkably increased compared to HD (figure 2G). Besides, frequencies of both IL-4^+^ CD8^+^ T cells (Tc2) and IL-17A^+^ CD8^+^ T cells (Tc17) in CR were significantly reduced (figure 2H and I). Moreover, Tc1, Tc2 and Tc17 in CR, consistently deviated from HD and showed no tendency to recover with the increased days post clinical discharge (figure 2E, H and I). The data suggested remarkable repression on CD8^+^ T cells of CR was long-lasting, though activation, proliferation and differentiation might be normal.

**Figure 2.**
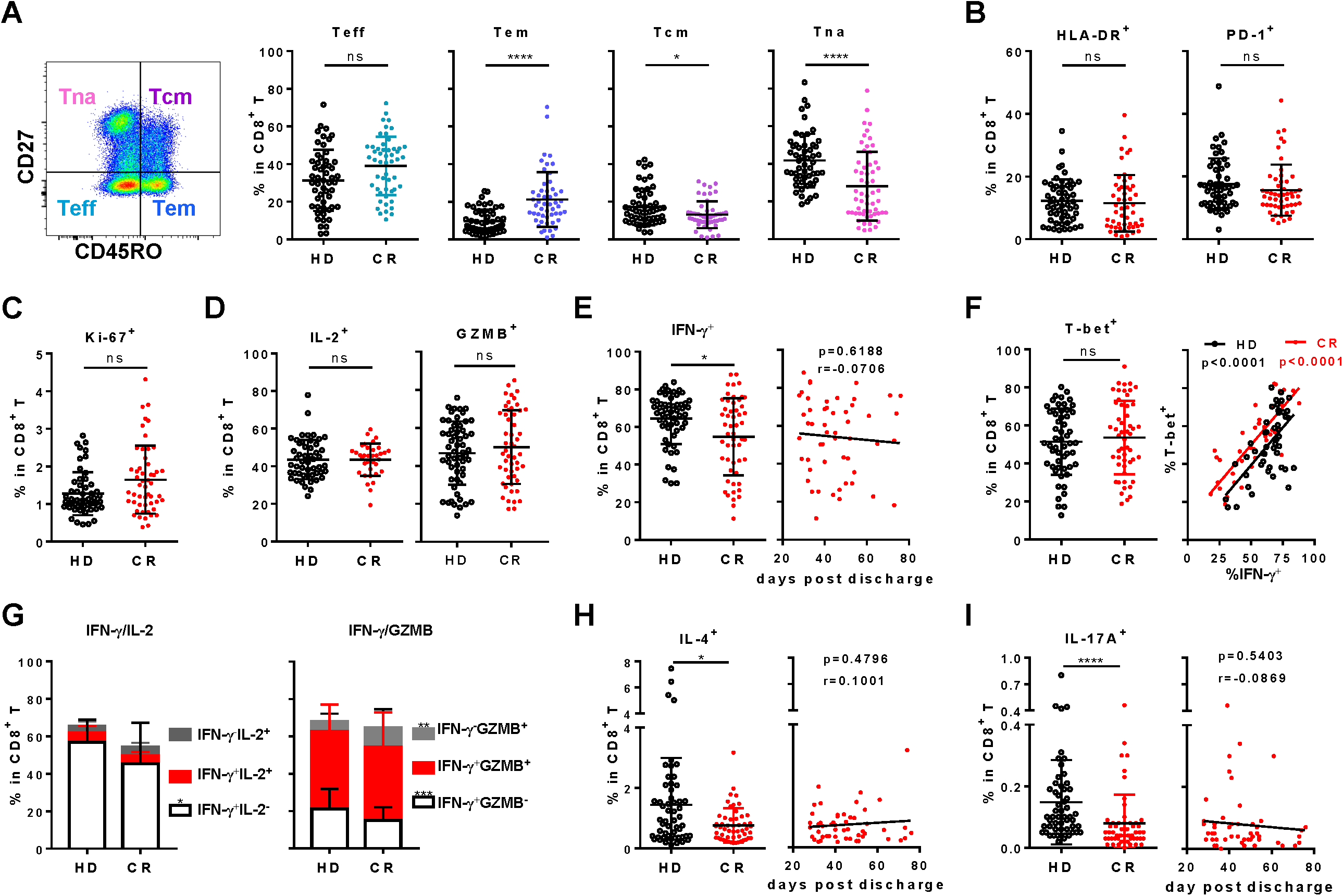
Differentiation, activation, proliferation and function of CD8^+^ T cells in peripheral blood of HD and CR. A, Gating strategy and the frequencies of effector T cells (Teff, CD45RO^-^CD27^-^), effector memory T cell (Tem, CD45RO^+^CD27^-^), central memory T cell (Tcm, CD45RO^+^CD27^+^) and naïve T cell (CD45RO ^-^CD27^+^, Tna) subsets in CD8^+^ T cells of PBMC. B and C, Frequencies of HLA-DR^+^, PD-1^+^ and Ki-67^+^ in CD8^+^ T cells. To analyze the function of CD8^+^ T cells, PBMCs were stimulated by PMA/Ionomycin for 4.5 h in the presence of BFA and monensin. The production of IL-2, Granzyme B (GZMB, D), IFN-γ (E), IFN-γ/IL-2, IFN-γ/GZMB co-expression (G), IL-4 (H) and IL-17A (I) by CD8^+^ T cells were analyzed by intracellular staining. F, T-bet^+^ cells in CD8^+^ T cells without stimulation (left panel), and correlation of T-bet^+^ with IFN-γ^+^ (right panel). The correlation of days post discharge in CR with frequency of IFN-γ^+^ (E), IL-4^+^ (H) and IL-17A^+^ (I) in CD8^+^ T cells were displayed in the right panels. ns, non-significant; *, P<0.05; **, P<0.01; ***, P<0.001; ****, P<0.0001.

In the context of CD4^+^ T cells, the frequency of Teff, Tem or naïve T cells of CR cohort showed no change, but Tcm was remarkably reduced compared to HD (figure 3A), suggesting an aberrant CD4^+^ T cell differentiation post SARS-CoV-2 infection. In terms of activation, HLA-DR on CD4^+^ T cells of CR and HD also showed no difference, though HLA-DR^+^ in naïve CD4 ^+^ T cells of CR was significantly lower (figure 3B and S2A). However, PD-1^+^ was remarkably lower in all CD4^+^ T cell subsets of CR (figure 3B and S2B), indicating a significant repression of the CD4^+^ T cell activation in CR. The proliferation indicated by Ki-67 showed no difference between CR and HD (figure 3C) though Ki-67^+^ in Tcm subset of CR was slightly higher (figure S2C). Function analysis under polyclonal stimulation showed the frequency of IFN-γ^+^ in CD4^+^ T cells (Th1) of CR was notably lower than that of HD (figure 3E), though the frequencies of IL-2^+^ and GZMB^+^ CD4^+^ T cell showed no difference between CR and HD (figure 3D). Similar as CD8^+^ T cells, T-bet in CD4^+^ T cells of CR was not reduced either and the correlation between T-bet^+^ and IFN-γ^+^ also remained (figure 3F). In more detail, the proportions of IFN-γ^+^ IL-2^+^ and IFN-γ^+^GZMB^-^ of CR were significantly reduced, but IFN-γ^-^GZMB^+^ remarkably increased in CD4^+^ T cells of CR (figure 3G). However, the frequencies of IL-4^+^ CD4^+^ T cell (Th2) and IL-17A^+^ CD4^+^ T cell (Th17) of CR (figure 3H and I) were both reduced significantly, indicating the functions of Th2 and Th17 were somehow repressed. Percentages of CXCR3^+^ and CCR6^+^, which could present Th1 and Th17 cells respectively, were also highly reduced in CD4^+^ T cells of CR (figure S2D and 2E), indicating the frequencies of Th1 and Th17 were persistently and remarkably reduced in CR.

**Figure 3.**
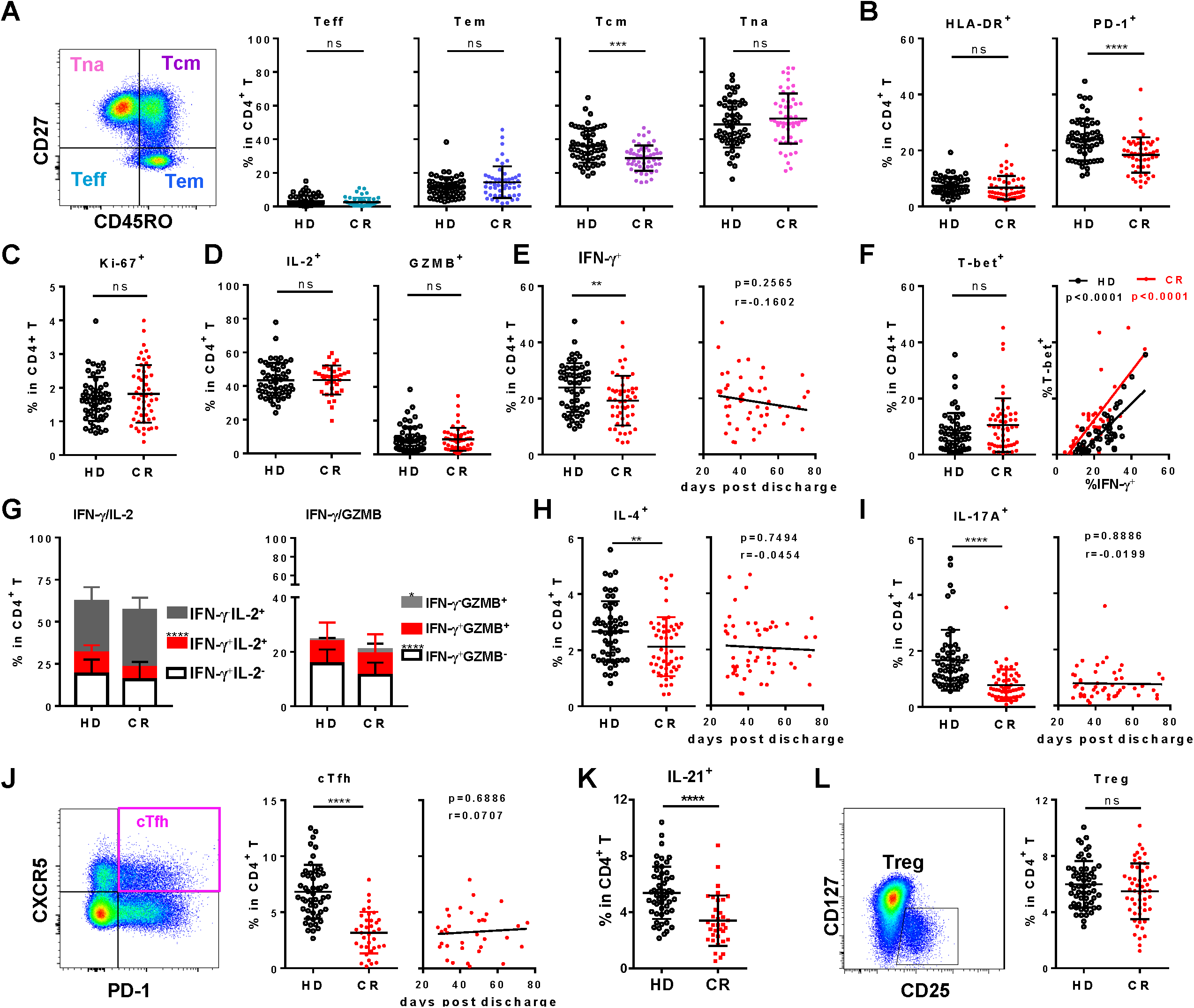
Differentiation, activation, proliferation and function of CD4^+^ T cells in peripheral blood of HD and CR. A, Frequencies of effector T cells (Teff, CD45RO^-^CD27^-^), effector memory T cell (Tem, CD45RO^+^CD27^-^), central memory T cell (Tcm, CD45RO^+^CD27^+^) and naïve T cell (CD45RO^-^CD27^+^, Tna) subsets in CD4^+^ T cells of PBMC. B and C, Frequencies of HLA-DR^+^, PD-1^+^ (B) and Ki-67^+^ (C) in CD4^+^ T cells. To analyze the function of CD4^+^ T cells, PBMCs were stimulated by PMA/Ionomycin for 4.5 h in the presence of BFA and monensin. The production of IL-2, GZMB (D), IFN-γ (E left panel), IFN-γ/IL-2, IFN-γ/GZMB co-expression (G), IL-4 (H), IL-17A (I) and IL-21 (K) by CD4^+^ T cells of HD and CR were analyzed by intracellular staining. F, T-bet^+^ cells in CD4^+^ T cells without stimulation (left panel), and correlation of T-bet^+^ with IFN-γ^+^ (right panel). J and L, Gating strategy and the frequency of circulating follicular helper T cells (cTfh, CXCR5^+^PD-1^+^) and Tregs (CD25^+^CD127^+^) in CD4^+^ T cells of HD and CR. The correlation of days post discharge in CR with frequency of IFN-γ^+^ (E), IL-4^+^ (H), IL-17A^+^ (I) and cTfh (J) in CD4^+^ T cells were displayed in the right panels. ns, non-significant; *, P<0.05; **, P<0.01; ****, P<0.0001.

T follicular helper (Tfh), with CXCR5 as a marker, is the specialized subset of CD4^+^ T cells needed for germinal centers and related B cell responses.^15^ In line with the frequency of CXCR3^+^ and CCR6^+^, that of CXCR5^+^ in CD4^+^ T cells of CR was also much lower than that of HD (figure S2F). Accordingly, the frequency of circulating Tfh (cTfh, PD-1^+^CXCR5^+^) in peripheral blood of CR were distinguishably lower than that in HD and the percentage of IL-21-expressing CD4^+^ T cells in CR was also lower than that in HD (figure 3J and K). cTfh cells could be further classified into three subsets with different capability: cTfh1 (CCR6^-^CXCR3^+^), cTfh2 (CCR6^-^CXCR3^-^) and cTfh17 (CCR6^+^CXCR3^-^).^16^ As same as their parent population, all of the three subsets were reduced significantly in the CR compared to HD (figure S3A). Similar to the whole CD4^+^ T cells, activation indicated by ICOS and proliferation indicated by Ki-67 in Tfh cells showed no differences between CR and HD (figure S3B and C). Moreover, Th1, Th2, Th17 and Tfh in CR, consistently deviated from HD and showed no tendency to recover with the increased days post clinical discharge (figure 3E, H, I and J).

In contrast to Th effector subsets, the regulatory T cells (CD3^+^CD8^-^CD4^+^CD127^-^CD25^+^, Treg) between CR and HD were not significantly different (figure 3L). As for Treg subsets, the frequency of activated Treg (CD45RA^-^FoxP3^hi^, aTreg) in CR was not different from that in HD though the resting Treg (CD45RA^+^FoxP3^lo^, rTreg) in CR showed a slight lower frequency (figure S4A). CTLA4 is one of the most important co-inhibitory molecules expressed by Tregs. As shown in Figure S4B, the frequencies of CTLA-4^+^ in Treg and rTreg in CR were similar as that in HD, except CTLA-4^+^ in aTreg of CR showed a relatively lower frequency.

Among the B cells, none of the frequencies of B cell subsets, antibody secreting cells (CD27^+^CD38^hi^, ASC), naïve B cell s (CD27^-^CD38^lo^) and memory B cells (CD27^+^CD38^lo^, MBCs), were different from that of HD (figure 4A). However, isotype-switched MBCs (IgM^-^CD20^hi^) in CR was significantly lower than that of HD (figure 4B). More detailed analysis on B cell activation marker CD71 and co-stimulatory molecule ICOSL were carried out. Frequencies of CD71^+^ in both IgM^+^ MBCs and IgM^-^ MBCs but not naïve B cells of CR were higher than that of HD (figure 4C). Frequencies of ICOSL^+^ in all IgM^+^ MBCs, IgM^-^MBCs, and naïve B cells of CR showed no significant difference with that of HD, while higher in some individuals (figure 4C). The SARS-CoV-2 infection imposed effect on isotype-switched MBCs also appeared long lasted (figure 4B), although MBCs in CR are still in the process of active proliferation.

**Figure 4.**
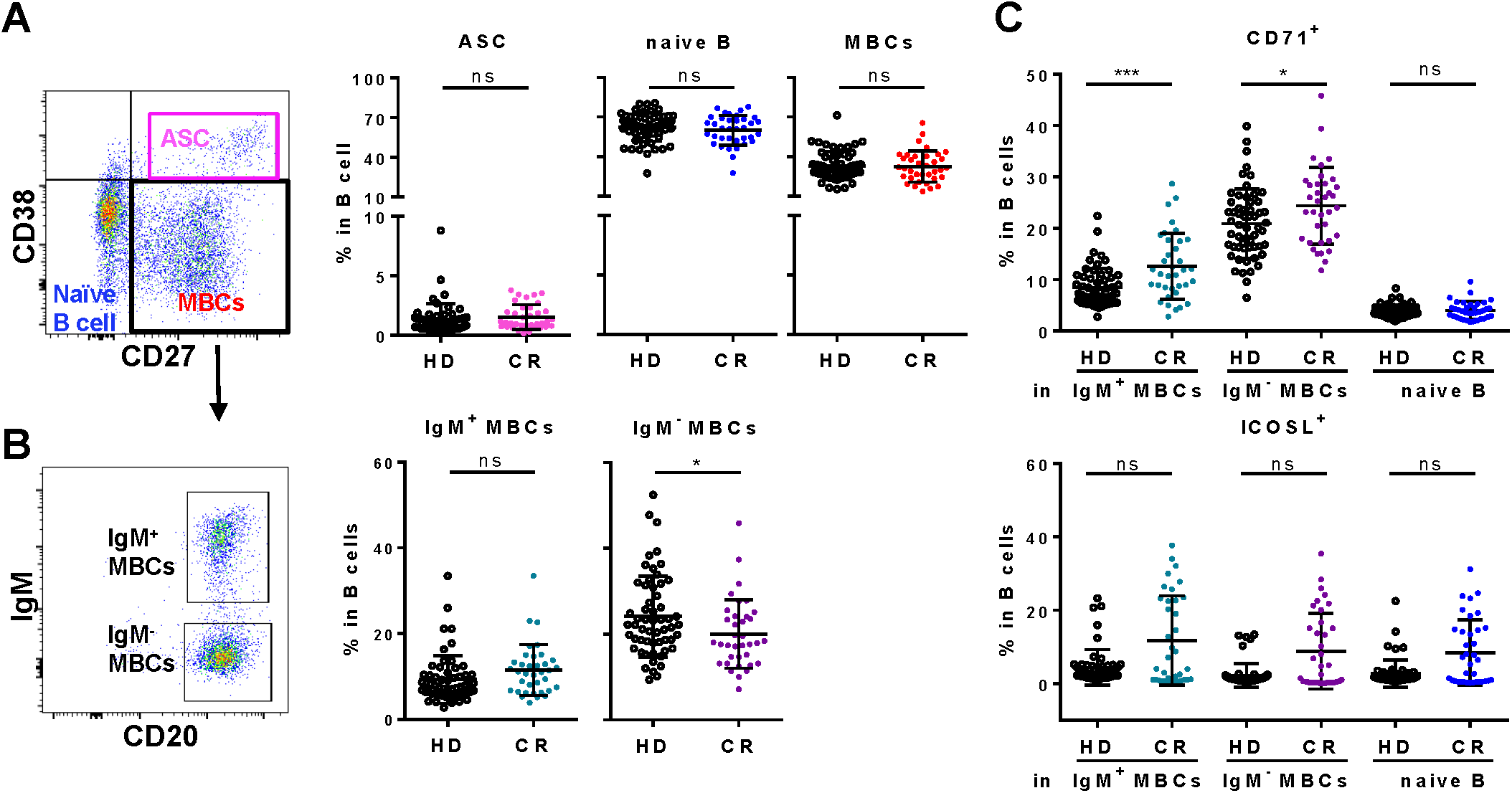
Differentiation and activation of B cells in peripheral blood of HD and CR. A, Gating strategy and the frequencies of naïve B cells (CD38 ^lo^CD27^-^), antigen secreting cell (ASC, CD38^hi^CD27^-^) and memory B cells (MBCs, CD38^lo^CD27^+^) subsets in B cells. B, The frequencies of non-isotype-switched IgM^+^ MBCs (IgM^+^CD20^hi^) and isotype-switched IgM^-^ MBCs (IgM^-^CD20^hi^) subsets in B cells. C, The frequencies of CD71^+^ and ICOS^+^ in IgM^+^ MBCs, IgM^-^ MBCs and naïve B cells. ns, non-significant; *, P<0.05.

In CD3^-^HLA-DR^lo^ of CR, the frequency of Ki-67^+^ was significantly lower than that of HD (figure 5A). Furthermore, under polyclonal stimulation, the frequency of GZMB^+^ and IFN-γ^+^ in CD3^-^HLA-DR^lo^ of CR were both significant lower than those of HD (figure 5B and C). Among the CD3^-^HLA-DR^lo^, the proportion of IFN-γ^+^GZMB^+^ cells in CR was remarkably reduced, while the proportion of IFN-γ^-^GZMB^+^ was markedly increased compared to those in HD (figure 5D). On the other hand, the frequency of T-bet expressing cells in CD3^-^HLA-DR^lo^ of CR was also reduced (figure 5E). Furthermore, correlation between T-bet^+^ and IFN-γ^+^ disappeared in CD3^-^HLA-DR^lo^ of CR, compared to the good correlation in that of HD. Further analysis showed that about 85% cells of the CD3^-^HLA-DR^lo^ population were CD56^+^ NK cells in both CR and HD cohort (figure S5A and B), suggesting the phenotypes of CD3^-^HLA-DR^lo^ shown above were mostly ascribed to NK.

**Figure 5.**
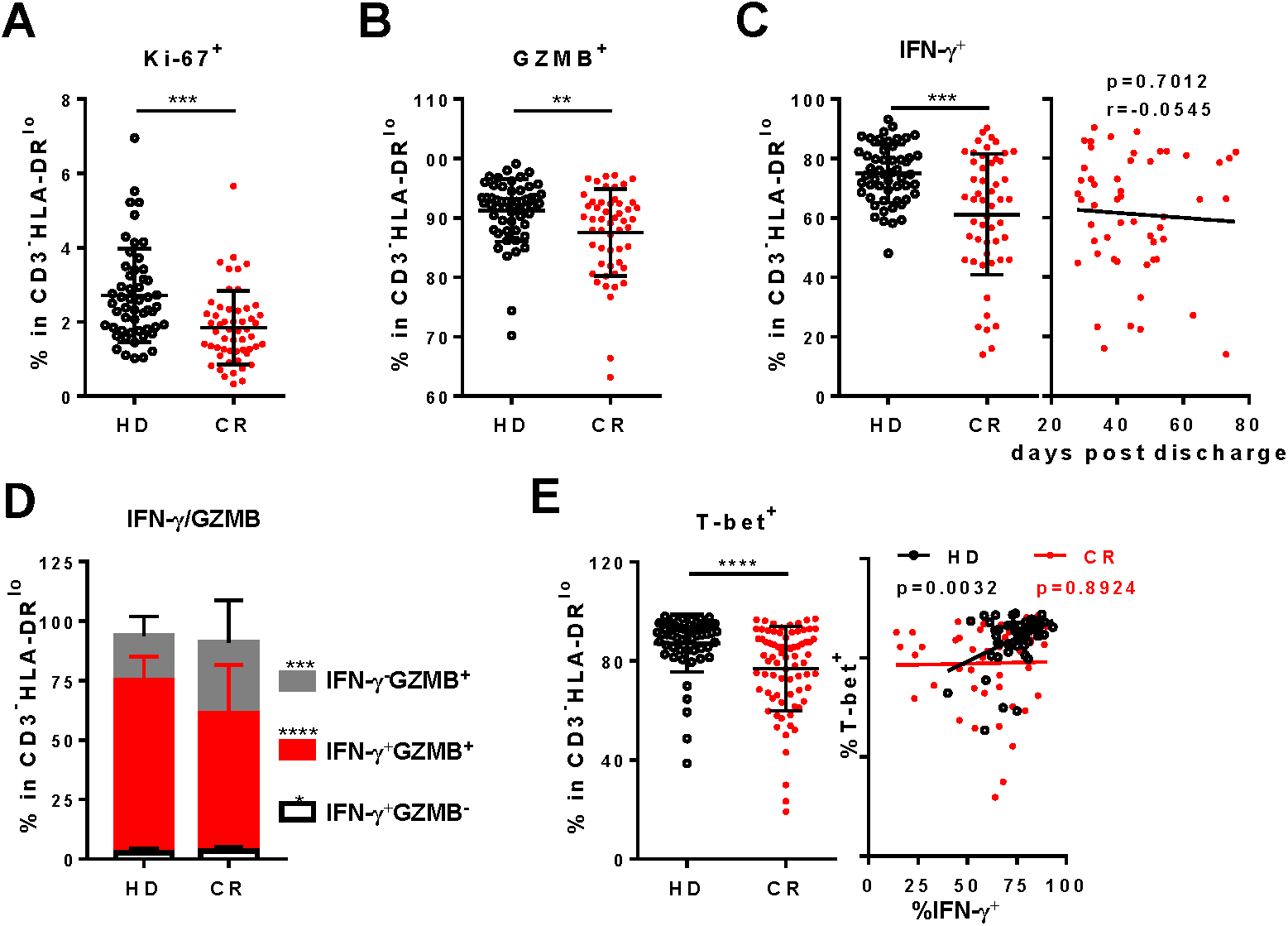
Proliferation and function of CD3^-^HLADR^lo^ lymphocytes cells in peripheral blood of HD and CR. A, Frequency of Ki-67^+^cells in CD3^-^HLADR^lo^ lymphocytes in PBMCs. To analyze the function of CD3^-^HLADR^lo^ lymphocytes, PBMCs were stimulated by PMA/Ionomycin for 4.5 h in the presence of BFA and monensin. The production of GZMB (B), IFN-γ (left panel of C) and IFN-γ/GZMB co-expression (D) by CD3^-^HLADR^lo^ lymphocytes were analyzed by intracellular staining. E, T-bet^+^ cells in CD3^-^HLADR^lo^ lymphocytes without stimulation (left panel) and correlation of T-bet^+^ with IFN-γ^+^ (right panel) in CD3^-^HLADR^lo^ lymphocytes. Correlation of days post discharge in CR with frequency of IFN-γ^+^ in CD3^-^HLADR^lo^ lymphocytes were displayed in the right panel of C. *, P<0.05; **, P<0.01; ***, P<0.001; ****, P<0.0001.

## Discussion

In this study, we generated a cross sectional lymphocyte response data of a clinical recovered COVID-19 cohort in Wuhan. By using flow cytometry with different molecular marker sets, we depicted an overview picture of CD8^+^ T, CD4^+^ T, B as well as CD3^-^HLA-DR^lo^ cell subsets on the aspects of differentiation, proliferation, activation and function. In CR individuals, a significant loss of CD4^+^ T cells still sustained long after clinical recovery from characteristic lymphopenia caused by acute SARS-CoV-2 infection. Specifically, the loss was embodied in significant degree on the decreased percentages of CD4^+^ Tcm in CD4^+^ T cell population (figure 1B) of CR cohort.

What’s even more remarkable is that profound lowered functions was observed in almost all T cell subsets we tested for the CR cohort, including Th1, Th2, Th17, Tfh, Tc1, Tc2, Tc17. The lowered functions were persistent to even 11 weeks after the CR cohort had clinically recovered. This suggests that the COVID-19 patients experienced long lasting repression on functions in general in both CD4^+^ and CD8^+^ T cells. The long-lasting dysfunction of T lymphocytes is common in chronic virus infected patients such as AIDS and hepatitis C, or cancer patients,^17-19^ but is rarely reported in acute virus infection, except the reported loss of Th17 in influenza infected individuals.^20^ To our knowledge, there is currently no report to tell whether this kind of long-lasting lowered function happens or not in the highly pathogenic corona viruses, MERS-CoV or SARS-CoV infected patients. Our findings in the present study suggested that the SARS-CoV-2 infection likely left unique imprints on lymphocytes and kept suppression on the functions of lymphocytes for a long time. The mechanism underlying the specific lymphocyte loss in COVID-19 patients warrant further investigations.

It should be noted that the frequencies of cTfhs were reduced significantly in the CR compared to HD, but the expression levels of both Ki-67 and ICOS in cTfh between CR and HD were similar, suggesting no suppression on the proliferation and activation of cTfh of CR (figure 3J and S3). This non-suppression is in line with the elevated activation indicated by CD71 and ICOSL in memory B cells of CR.^21^ The lower frequency of IgM^-^ isotype-switched MBC in CR than in HD, indicated the induction of isotype-switched MBC might be repressed by SARS-CoV-2 infection. This repression of isotype-switch might be related with the reduced frequency of cTfh, which are critical for the differentiation and isotype-switch of germ center B cells.^21^ In recent report on the antibody response of SARS-CoV-2 recovered individuals, the SARS-CoV-2 specific antibody responses sharply waned within 8 weeks.^22^ As the IgM^-^ switched MBC are more responsive than IgM^+^ MBC,^23^ the decrease of isotype-switched MBC might be one reason for explaining the sharp waning of SARS-CoV-2 specific antibody responses.

We further found that the proportions of IFN-γ^-^GZMB^+^ cells were significantly higher in CD4^+^ T cells, CD8^+^ T cells as well as in CD3^-^HLA-DR^lo^ lymphocytes of CR cohort compared to HD in contrary to the lower proportions of IFN-γ^+^ cells in the corresponding cell populations (figure 1D, 2D and 5E). In the CD3^-^HLA-DR^lo^ population, the correlation between T-bet and IFN-γ disappeared in CR cohort compared to the nice correlation in HD cohort (figure 5H). These data suggested perturbation also existed in T cells and CD3^-^HLA-DR^lo^ lymphocytes of COVID-19 CR individuals besides repression. However, further classification on the CD3^-^HLA-DR^lo^ lymphocytes was missed in this study due to lack of corresponding antibodies during the COVID-19 crisis in Wuhan. Although about 85% of the CD3^-^HLA-DR^lo^ lymphocytes might be NK as we assayed further in either CR or HD cohort, there is another important cell population, innate lymphocytes (ILCs), also located in CD3^-^HLA-DR^lo^. The responses and status of the NKs and ILCs in CR should be of high interest for understanding COVID-19.

In summary, we provided a cross sectional profile of lymphocyte responses of a clinical recovered COVID-19 cohort and found long-term significant phenotype alterations and potential dysfunctions of lymphocytes in the cohort clinically recovered from laboratory-confirmed COVID-19. We still do not know how long the phenotype alterations and potential dysfunctions of lymphocytes will last. As reported, CD4^+^ T cells and CD8^+^ T cells and NK cells are all critical for the control of intracellular pathogen infections and tumors^17,24^,25 and could coordinate with each other^.26,27^ CD4^+^ T cells, especially the Tfh, and B cells are also critical for the processes that lead to long-term humoral immunity^16^. The broad and long-term dysfunction of these lymphocytes subsets might profoundly impair the immune surveillance and protection executed by lymphocytes in the COVID-19 clinical recovered individuals, though broad and strong SARS-CoV-2 specific memory CD4^+^ T cells and CD8^+^ T cells could be detected in COVID-19 patients^.28,29^ A recent study on the SARS-CoV-2-specific humoral and cellular immunity in COVID-19 convalescent individuals highlighted that anti-viral T cells may not be maintained at high numbers in the PBMCs in the recovered patients.^30^ This alerts us more concerns on the prognosis of COVID-19 patients. Considering that SARS-CoV-2 specific antibody response also sharply waned,^22^ it is hard to speculate whether the SARS-CoV-2 clinical recovered population could resist re-infection for a long period. If the dysfunction of lymphocytes sustained long time, this population might be even more susceptible to SARS-CoV-2 infection or other viral infections. Hence, more comprehensive profile studies on the relationship of immune responses and SARS-CoV-2 infection longitudinally and cross-sectionally in different cohorts are urgently needed.

## Contributors

JY, MZ, EZ contributed to the conception, design, data acquisition, analysis, and interpretation, and drafted and critically revised the manuscript. KH contributed to the data acquisition, clinical descriptions and critically revised the manuscript. QY, DZ, JiaX, YC, MS, BZ, JieX, YL, YH contributed to the data acquisition and analysis. XZ, CH and YS made contribution to the study concept, design and critically revised the manuscript. HY contributed to the conception, design, data analysis, and interpretation, and drafted and critically revised the manuscript. All of the authors gave final approval and agreed to be accountable for all aspects of the work.

## Declaration of interests

We declare no competing interests.

## Data Availability

I can provide the data of this study if required.

## Acknowledgments

We thank the COVID-19 recovered individuals and healthy donors involved in this study and staff at Wuhan Jinyintan Hospital.

## Supplementary figures

**Figure S1.**
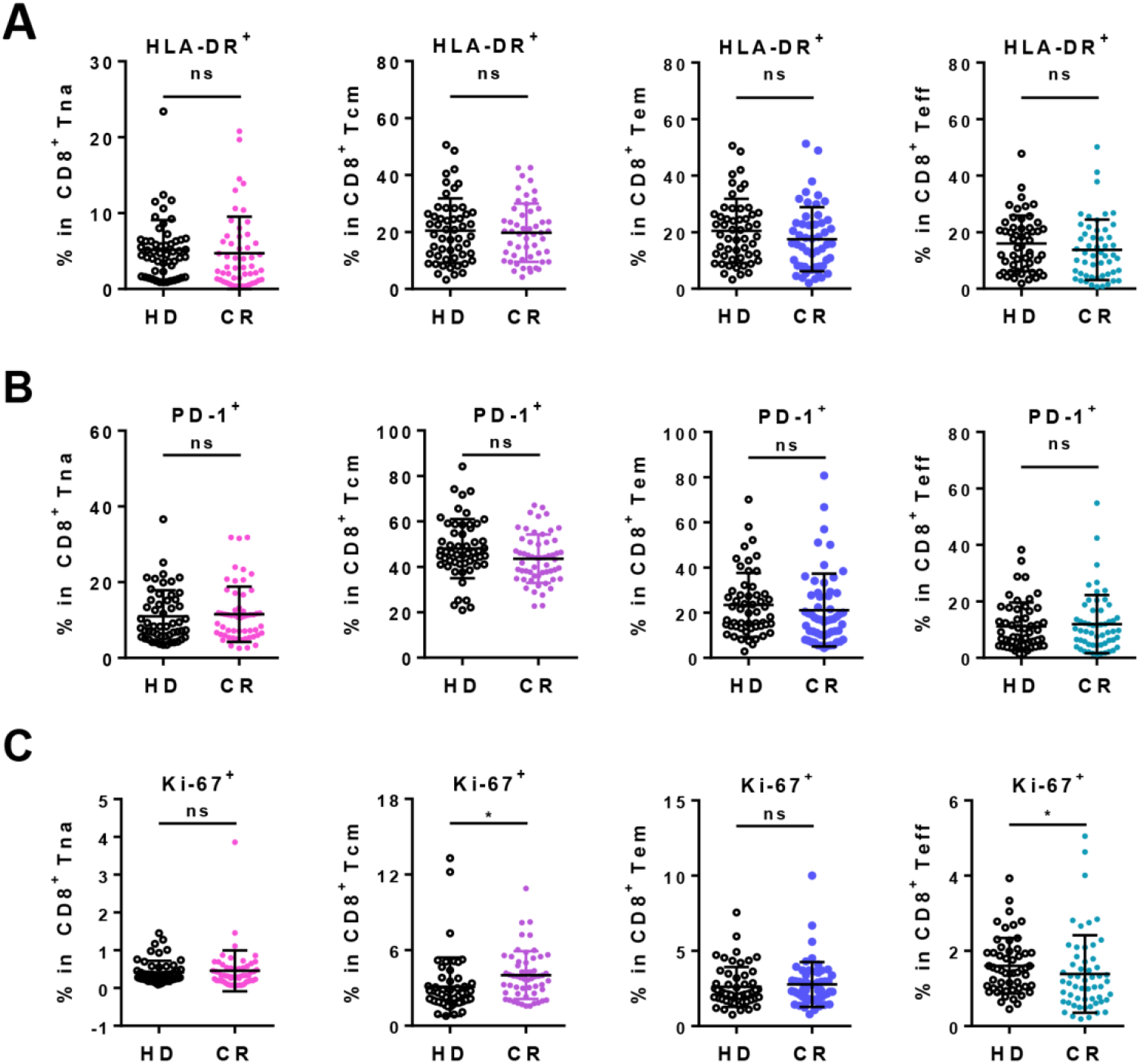
Activation and proliferation of the Tna, Tcm, Tem and Teff subsets of CD8^+^ T cells. Frequencies of HLA-DR^+^ (A), PD-1^+^ (B) and Ki-67^+^ (C) in naïve T cells (Tna), central memory T cells (Tcm), effector memory T cell (Tem) and effector T cell (Teff) subsets of CD8^+^ T cells in PBMCs of healthy donors (n=55) and clinically recovered COVID-19 individuals (CR). ns, non-significant; *, P<0.05.

**Figure S2.**
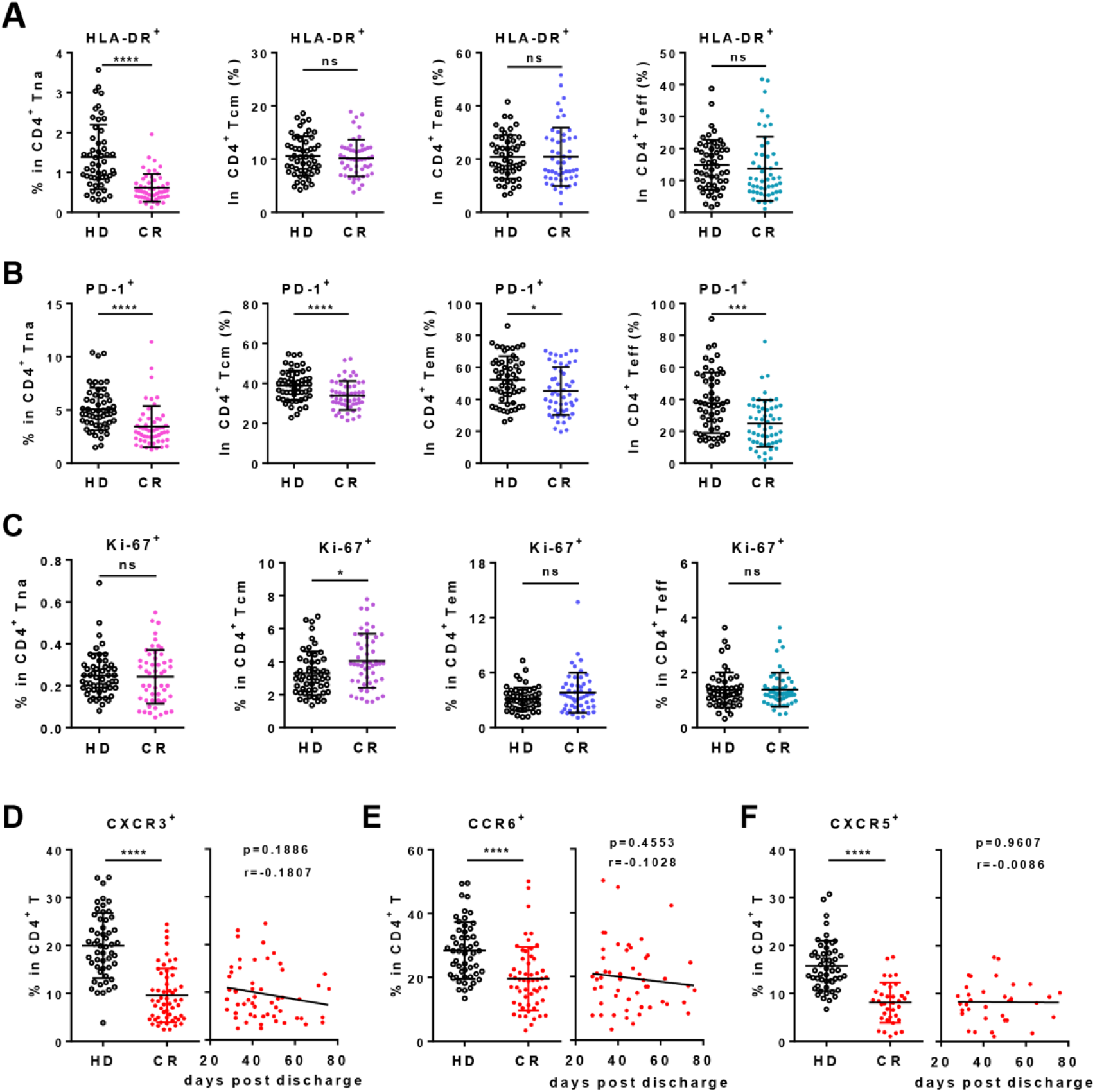
Activation and proliferation of the Tna, Tcm, Tem and Teff subsets of CD4^+^ T cells and chemokine receptor expression in CD4^+^ T cells. Frequencies of HLA-DR^+^ (A), PD-1^+^(B) and Ki-67^+^ (C) in naïve T cells (Tna), central memory T cells (Tcm), effector memory T cell (Tem) and effector T cell (Teff) subsets of CD4^+^ T cells in PBMCs of healthy donors (n=55) and clinically recovered COVID-19 individuals (CR). D-F, Frequencies of CXCR3^+^ (D), CCR6^+^ (E) and CXCR5^+^ (F) in CD4^+^ T cells were shown in the left panel of C-E. The correlation of days post discharge with frequency of CXCR3^+^ (D), CCR6^+^ (E) and CXCR5^+^ (F) in CD4^+^ T cells of CR were displayed in the right panel of C-E. ns, non-significant; *, P<0.05; ***, p<0.001; ****, p<0.0001.

**Figure S3.**
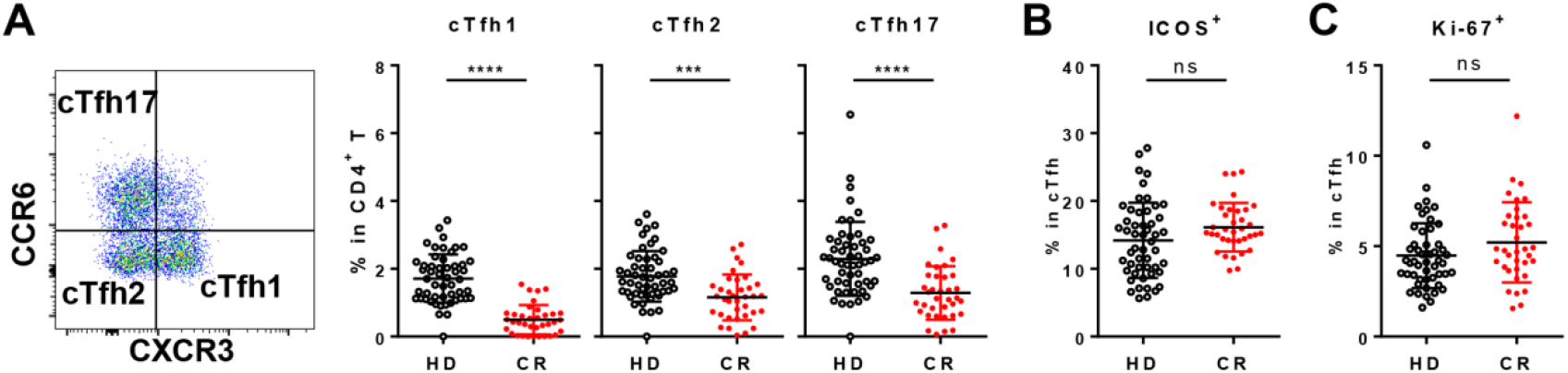
Frequencies of cTfh subsets and the activation, proliferation of cTfh in peripheral blood of HD and CR. A, Gating strategy and the frequencies of cTfh subsets cTfh1 (CXCR3^+^ CCR6^-^), cTfh2 (CXCR3^-^ CCR6^-^) and cTfh17 (CXCR3^-^ CCR6^+^) of CD4^+^ T cells in PBMCs of healthy donors (n=55) and clinically recovered COVID-19 individuals (CR). B and C, The frequencies of ICOS^+^ and Ki-67^+^cells in cTfh. ns, non-significant; ***, p<0.001; ****, p<0.0001.

**Figure S4.**
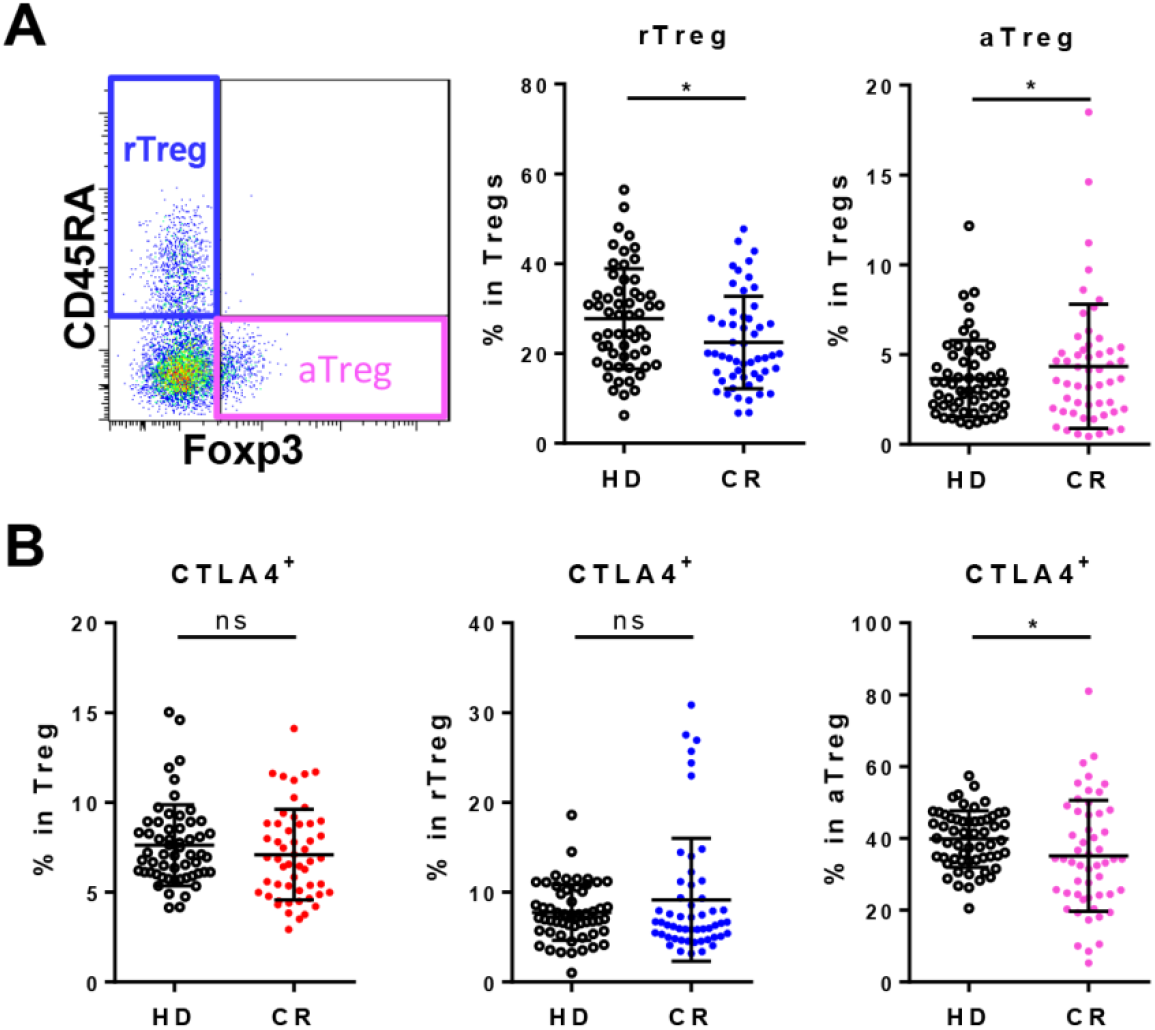
Frequencies and phenotype of Tregs and Treg subsets in peripheral blood of HD and CR. A, Gating strategy and the frequencies of resting Treg (rTeg, CD45RA^+^Foxp3^lo^) and activated Treg (aTreg, CD45RA^-^Foxp3^hi^) in Treg. B, The frequencies of CTLA-4^+^ cells in Treg, rTreg and aTreg. ns, non-significant; *, P<0.05.

**Figure S5.**
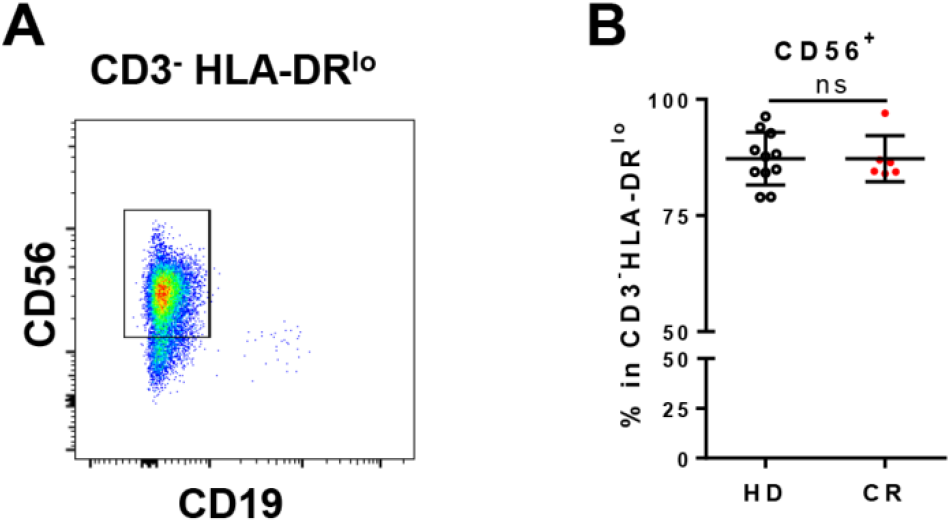
Frequencies of CD56^+^ NK cells in CD3^-^HLADR^lo^ lymphocytes cells of peripheral blood. Gating strategy and the frequencies of CD56^+^ NK cells (CD56^+^CD19^-^) in CD3^-^HLADR^lo^ lymphocytes of PBMCs derived from HD (n=8) and CR (n=5). ns, non-significant.

## Supplementary tables

**Supplementary Table 1.** (Table S1): clinical information of CR cohort.

| final code | sex | age | day of discharge | day of testing | days post discharge | SARS-CoV-2 antibody |  | blood oxygen saturation | symptoms in two weeks |  |  |  |  | blood routine examination (10<sup>9</sup>/L) |  |  |
| --- | --- | --- | --- | --- | --- | --- | --- | --- | --- | --- | --- | --- | --- | --- | --- | --- |
|  |  |  |  |  |  | IgG | IgM |  | fever | shod of breath | cough | expectoration | oxygen uptake | WBC | neutrophils | lymphocytes |
| 1 | M | 43 | 2020-3-17 | 2020-4-14 | 28 | + | + | 99 | - | - | - | - | - | 6.22 | 3.81 | 1.95 |
| 2 | F | 46 | 2020-2-5 | 2020-4-14 | 44 | + | + | 99 | - | - | + | - | - | 6.52 | 1.59 | 1.8 |
| 3 | F | 50 | 2020-3-13 | 2020-4-14 | 32 | + | - | 99 | - | - | - | - | - | 5.54 | 3.66 | 1.33 |
| 4 | M | 68 | 2020-3-12 | 2020-4-14 | 39 | + | - | 99 | - | - | - | - | - | 5.68 | 3.47 | 1.91 |
| 5 | F | 40 | 2020-3-1 | 2020-4-14 | 54 | + | + | 99 | - | - | - | - | - | 5.21 | 3.88 | 1.03 |
| 6 | F | 48 | 2020-3-2 | 2020-4-14 | 73 | + | - | 99 | - | - | - | - | - | 6.06 | 2.71 | 2.94 |
| 7 | F | 39 | 2020-3-2 | 2020-4-14 | 45 | + | + | 98 | - | - | - | - | - | 6.26 | 4.12 | 1.64 |
| 8 | M | 45 | 2020-3-13 | 2020-4-14 | 61 | + | + | 99 | - | - | - | - | - | 5.06 | 2.64 | 1.81 |
| 9 | M | 60 | 2020-2-28 | 2020-4-14 | 41 | + | - | 98 | - | - | - | - | - | 3.75 | 2.12 | 1.26 |
| 10 | F | 34 | 2020-3-6 | 2020-4-14 | 41 | + | - | 98 | - | - | - | - | - | 5.55 | 3.06 | 1.88 |
| 11 | M | 32 | 2020-2-20 | 2020-4-14 | 29 | + | - | 95 | - | - | - | - | - | 6.56 | 4.12 | 2.07 |
| 12 | M | 55 | 2020-1-31 | 2020-4-14 | 32 | + | - | 96 | - | - | - | - | - | 5.33 | 1.9 | 3.01 |
| 13 | F | 54 | 2020-2-1 | 2020-4-14 | 38 | + | - | 99 | - | - | - | - | - | 4.78 | 2.85 | 1.48 |
| 14 | M | 35 | 2020-3-5 | 2020-4-14 | 40 | + | + | 98 | - | - | - | - | - | 6.9 | 3.44 | 2.95 |
| 15 | F | 57 | 2020-2-29 | 2020-4-14 | 41 | + | - | 99 | - | + | - | - | - | 5.69 | 3.15 | 2.01 |
| 16 | F | 27 | 2020-3-10 | 2020-4-14 | 44 | + | - | 99 | - | - | - | - | - | 5.29 | 2.7 | 2.2 |
| 17 | F | 31 | 2020-2-13 | 2020-4-14 | 37 | + | + | 99 | - | - | - | - | - | 6.36 | 3.18 | 2.32 |
| 18 | F | 51 | 2020-3-14 | 2020-4-14 | 40 | + | + | 99 | - | - | - | - | - | 3.95 | 2.22 | 1.41 |
| 19 | F | 34 | 2020-3-12 | 2020-4-14 | 51 | + | + | 98 | - | - | - | - | - | 7.22 | 4.46 | 2.21 |
| 20 | F | 68 | 2020-2-14 | 2020-4-14 | 74 | + | + | 99 | - | - | - | - | - | 4.08 | 3.22 | 0.64 |
| 21 | M | 60 | 2020-3-15 | 2020-4-14 | 47 | + | + | 99 | - | - | - | - | - | 6.31 | 3.76 | 2.01 |
| 22 | F | 31 | 2020-3-17 | 2020-4-14 | 55 | + | + | 98 | - | - | - | - | - | 7.24 | 3.45 | 3.12 |
| 23 | F | 39 | 2020-2-12 | 2020-4-14 | 73 | + | + | 96 | - | - | - | - | - | 6.09 | 3.56 | 2.06 |
| 24 | M | 37 | 2020-2-22 | 2020-4-15 | 33 | + | - | 99 | - | - | - | - | - | 6.96 | 4.78 | 1.76 |
| 25 | M | 70 | 2020-3-4 | 2020-4-15 | 34 | + | - | 98 | - | + | - | - | - | 7.13 | 4.38 | 2.08 |
| 26 | F | 51 | 2020-3-18 | 2020-4-15 | 44 | + | - | 99 | - | - | - | + | - | 7.98 | 5 | 2.44 |
| 27 | F | 40 | 2020-3-1 | 2020-4-15 | 46 | + | - | 99 | - | - | - | - | - | 5.17 | 3.13 | 1.66 |
| 28 | F | 51 | 2020-3-4 | 2020-4-15 | 41 | + | - | 99 | - | - | - | - | - | 3.82 | 1.92 | 1.42 |
| 29 | F | 61 | 2020-2-22 | 2020-4-15 | 36 | + | + | 98 | - | - | - | - | - | 6.85 | 3.74 | 2.39 |
| 30 | F | 67 | 2020-3-14 | 2020-4-15 | 32 | + | - | 98 | - | + | - | - | - | 7.32 | 3.68 | 2.9 |
| 31 | M | 56 | 2020-3-18 | 2020-4-15 | 34 | + | + | 99 | - | - | - | - | - | 7.96 | 4.81 | 2.5 |
| 32 | M | 51 | 2020-3-16 | 2020-4-15 | 63 | + | - | 98 | - | - | - | - | - | 8.58 | 6.74 | 1.38 |
| 33 | F | 61 | 2020-3-6 | 2020-4-15 | 28 | + | + | 98 | - | - | - | - | - | 6.13 | 3.43 | 2.15 |
| 34 | M | 31 | 2020-2-25 | 2020-4-15 | 53 | + | - | 98 | - | - | + | - | - | 6.38 | 4.06 | 2.01 |
| 35 | M | 53 | 2020-2-10 | 2020-4-15 | 40 | + | + | 99 | - | - | - | - | - | 5.19 | 2.86 | 2 |
| 36 | F | 38 | 2020-3-13 | 2020-4-15 | 50 | + | - | 99 | - | - | - | - | - | 5.02 | 2.78 | 1.87 |
| 37 | F | 40 | 2020-3-7 | 2020-4-15 | 65 | + | - | 99 | - | - | - | - | - | 5.49 | 2.8 | 2.33 |
| 38 | M | 54 | 2020-3-5 | 2020-4-15 | 51 | + | - | 96 | - | - | - | - | - | 5.15 | 2.79 | 1.9 |
| 39 | F | 56 | 2020-2-24 | 2020-4-15 | 33 | + | - | 98 | - | - | - | - | + | 6.11 | 3.43 | 1.97 |
| 40 | F | 38 | 2020-3-4 | 2020-4-15 | 49 | + | + | 99 | - | - | - | - | - | 4.88 | 2.38 | 2.07 |
| 41 | F | 43 | 2020-3-1 | 2020-4-15 | 52 | + | - | 99 | - | - | - | - | - | 8.86 | 5.14 | 3.09 |
| 42 | F | 57 | 2020-2-23 | 2020-4-16 | 71 | + | + | 99 | - | - | + | - | - | 6.02 | 3.67 | 1.94 |
| 43 | M | 34 | 2020-3-13 | 2020-4-16 | 45 | + | - | 99 | - | - | + | - | - | 3.97 | 2.06 | 1.6 |
| 44 | F | 61 | 2020-3-8 | 2020-4-16 | 76 | + | - | 99 | - | - | + | + | - | 4.98 | 3.24 | 1.4 |
| 45 | F | 57 | 2020-3-5 | 2020-4-16 | 62 | + | - | 99 | - | - | - | - | - | 6.63 | 4.71 | 1.45 |
| 46 | M | 55 | 2020-2-23 | 2020-4-16 | 32 | + | - | 96 | - | + | + | + | - | 7.17 | 5.34 | 1.37 |
| 47 | M | 51 | 2020-2-26 | 2020-4-16 | 30 | + | - | 99 | - | - | - | - | - | 4.56 | 2.46 | 1.54 |
| 48 | M | 38 | 2020-3-14 | 2020-4-16 | 54 | + | - | 99 | - | - | - | - | - | 6.1 | 3.8 | 1.82 |
| 49 | F | 51 | 2020-2-26 | 2020-4-16 | 46 | + | + | 99 | - | - | - | - | - | 4.88 | 2.82 | 1.54 |
| 50 | M | 56 | 2020-3-17 | 2020-4-16 | 33 | + | - | 96 | - | - | - | - | - | 5.24 | 2.17 | 2.63 |
| 51 | M | 61 | 2020-1-31 | 2020-4-16 | 29 | + | + | 99 | - | - | - | - | - | 3.93 | 1.83 | 1.75 |
| 52 | F | 39 | 2020-2-27 | 2020-4-16 | 53 | + | - | 99 | - | - | - | - | - | 5.32 | 4.94 | 3.61 |
| 53 | F | 52 | 2020-2-19 | 2020-4-16 | 50 | + | - | 99 | - | - | - | - | - | 6.66 | 3.67 | 2.56 |
| 54 | F | 70 | 2020-2-1 | 2020-4-16 | 33 | + | - | 99 | - | - | - | - | - | 5.57 | 3.18 | 1.9 |
| 55 | F | 65 | 2020-2-23 | 2020-4-16 | 30 | + | - | 99 | - | - | - | - | - | 5.46 | 2.94 | 2.1 |

+, positive; -, negative.
Cells filled in yellow color means the value was higher than normal range while cells filled in blue color means the value was lower than normal range

**Supplementary Table 2.** (Table S2): reagents for flow cytometry.

| antibodies (clone) | staining procedure | source | identifier |
| --- | --- | --- | --- |
| Fixable Viability Dye eFluor™ 506 | surface | eBioscience | 65-0866-14 |
| Alexa Fluor® 700 anti-human CD183 (CXCR3) (G025H7) | surface | BioLegend | 353742 |
| Alexa Fluor® 700 anti-human CD3 (SK7) | surface | BioLegend | 344822 |
| Alexa Fluor® 700 anti-human CD4 (OKT4) | surface | BioLegend | 317426 |
| APC anti-human CD196 (CCR6) (G034E3) | surface | BioLegend | 353416 |
| APC anti-human CD275 (B7-H2, ICOSL) (2D3) | surface | BioLegend | 309408 |
| APC/Cyanine7 anti-human CD3 (HIT3a) | surface | BioLegend | 300318 |
| APC/Cyanine7 anti-human CD45RA (HI100) | surface | BioLegend | 304127 |
| Brilliant Violet 421™ anti-human/mouse/rat CD278 (ICOS)(C398.4A) | surface | BioLegend | 313524 |
| Brilliant Violet 605™ anti-human CD19 (HIB19) | surface | BioLegend | 302244 |
| Brilliant Violet 605™ anti-human CD8a (RPA-T8) | surface | BioLegend | 301040 |
| Brilliant Violet 650™ anti-human CD38 (HB-7) | surface | BioLegend | 356620 |
| Brilliant Violet 650™ anti-human CD4 (RPA-T4) | surface | BioLegend | 300536 |
| FITC anti-human CD20 (2H7) | surface | BioLegend | 302304 |
| FITC anti-human CD25 (BC96) | surface | BioLegend | 302604 |
| FITC anti-human CD45RO (UCHL1) | surface | BioLegend | 304242 |
| Pacific Blue™ anti-human CD27 ( O323) | surface | BioLegend | 302822 |
| PE anti-human IgM (MHM-88) | surface | BioLegend | 314508 |
| PE/Cy7 anti-human CTLA-4 (L3D10) | surface | BioLegend | 349914 |
| PE/Cyanine7 anti-human CD27 (O323) | surface | BioLegend | 302838 |
| PE/Dazzle™ 594 anti-human CD279 (PD-1 (EH12.2H7) | surface | BioLegend | 329940 |
| PE/Dazzle™ 594 anti-human CD71 (CY1G4) | surface | BioLegend | 334120 |
| PerCP/Cy5.5 anti-human CD127 (A019D5) | surface | BioLegend | 351322 |
| PerCP/Cyanine5.5 anti-human CD185 (CXCR5) (J252D4) | surface | BioLegend | 356910 |
| PerCP/Cyanine5.5 anti-human HLA-DR (L243) | surface | BioLegend | 307630 |
| Brilliant Violet 421™ anti-human FoxP3 (206D) | intracellular | BioLegend | 320124 |
| Brilliant Violet 650™ anti-human IL-2 (MQ1-17H12) | intracellular | BioLegend | 500334 |
| FITC anti-human/mouse Granzyme B (QA16A02) | intracellular | BioLegend | 372206 |
| PE anti-human IL-21 (3A3-N2) | intracellular | BioLegend | 513004 |
| PE anti-human Ki-67 (Ki-67) | intracellular | BioLegend | 350504 |
| PE/Cy7 anti-human IL-4 (MP4-25D2 ) | intracellular | BioLegend | 500824 |
| PE/Cyanine7 anti-human Ki-67 ( Ki-67) | intracellular | BioLegend | 350526 |
| PE/Dazzle™ 594 anti-human IFN-γ (B27) | intracellular | BioLegend | 506530 |
| PerCP/Cy5.5 anti-human IL-17A (BL168) | intracellular | BioLegend | 512314 |

## Notes

### Competing Interest Statement

The authors have declared no competing interest.

### Author Declarations

This study was reviewed and approved by the Medical Ethical Committee of Wuhan Jinyintan hospital (approval number KY-2020-47.01).

